# Trastuzumab Maintenance Every 4 Weeks versus Every 3 Weeks in HER2-Positive Breast Cancer: A Retrospective Analysis of Long-term Outcomes

**DOI:** 10.64898/2025.12.02.25341297

**Authors:** Swaroop Revannasiddaiah, Irappa Madabhavi, Siddharth Vats, Mukesh Sharma, Priyanka Thakur, Kailash Chandra Pandey

## Abstract

**Background:** Standard trastuzumab maintenance therapy is administered every 3 weeks at 6mg/kg following completion of chemotherapy. However, geographic barriers and patient compliance issues in montane terrain settings may limit adherence to this schedule. We evaluated the efficacy and safety of alternative 4-weekly dosing (8mg/kg) compared to standard 3-weekly dosing in HER2-positive breast cancer patients.

**Methods:** This retrospective cohort study analyzed patients with HER2-positive breast cancer treated between June 2015 and January 2018 at centers serving mountainous regions. All patients received standard TCH (docetaxel, carboplatin, trastuzumab) induction therapy. For maintenance, patients chose either 3-weekly trastuzumab (6mg/kg, n=47) or 4-weekly trastuzumab (8mg/kg, n=34) based on geographic accessibility. Primary endpoints were disease-free survival (DFS) and overall survival (OS) analyzed using Kaplan-Meier methodology and log-rank tests at median 8-year follow-up.

**Results:** Eighty-one patients were analyzed (Stage I: 17.3%, Stage II: 22.2%, Stage III: 59.3%, Oligometastatic: 1.2%). Baseline characteristics were balanced between groups (p=0.665). At median 8-year follow-up, Kaplan-Meier analysis showed non-inferior survival outcomes for 4-weekly dosing. Neither regimen reached median survival.

Five-year disease-free survival was 68.1% vs 70.6% (log-rank p=0.726, HR 1.15, 95%CI 0.53-2.51) and five-year overall survival was 74.5% vs 79.4% (log-rank p=0.721, HR 1.17, 95%CI 0.49-2.79) for 3-weekly vs 4-weekly groups, respectively. Event rates were 16/47 (34.0%) vs 10/34 (29.4%) for DFS and 13/47 (27.7%) vs 8/34 (23.5%) for OS. Stage-specific analysis showed consistent trends across all stages.

**Conclusions:** Trastuzumab maintenance at 8mg/kg every 4 weeks demonstrates non-inferiority to standard 3-weekly dosing with numerical trends favoring the alternative schedule. Kaplan-Meier survival analysis with robust statistical methodology confirms equivalent long-term outcomes. This approach offers a viable option for patients facing geographic or logistical barriers to standard dosing, potentially improving treatment adherence while maintaining efficacy. Further prospective validation is warranted.

## INTRODUCTION

Trastuzumab, a humanized monoclonal antibody targeting the HER2 receptor, has revolutionized the management of HER2-positive breast cancer since its introduction. The standard maintenance dosing regimen of trastuzumab at 6mg/kg every three weeks, established through pivotal clinical trials, has become the global standard of care for adjuvant therapy following initial chemotherapy.[1][2] This dosing schedule was primarily chosen to align with concurrent chemotherapy administration rather than being optimized for trastuzumab’s unique pharmacokinetic properties.[3]

The pharmacokinetic profile of trastuzumab supports the exploration of alternative dosing strategies. With a terminal half-life of approximately 28 days and low systemic clearance, trastuzumab exhibits favorable characteristics for extended dosing intervals.[4][5] Recent pharmacokinetic modeling studies demonstrate that alternative schedules, including 4-weekly administration at higher doses, can maintain therapeutic trough concentrations above minimum effective levels while potentially reducing treatment burden.[6][7]

Geographic and socioeconomic barriers significantly impact patient access to optimal cancer care, particularly in resource-constrained settings.[8][9] Patients residing in rural or mountainous regions face substantial challenges including extended travel distances, loss of income, and disruption to family routines when adhering to standard 3-weekly treatment schedules.[10][11] These barriers can lead to treatment delays, reduced adherence, and ultimately compromised clinical outcomes.[12]

The concept of treatment optimization in oncology increasingly recognizes the need to balance clinical efficacy with practical considerations of patient access and quality of life.[13] Alternative dosing strategies that maintain therapeutic equivalence while improving treatment feasibility represent an important avenue for addressing healthcare disparities.[14] Previous studies have explored duration modifications, with trials comparing 6-month versus 12-month trastuzumab therapy, but limited data exist on frequency modifications for maintenance dosing.[15][16]

Our study addresses this knowledge gap by evaluating the real-world effectiveness of 4-weekly trastuzumab maintenance dosing (8mg/kg) compared to standard 3-weekly dosing (6mg/kg) in a challenging geographic setting. The primary objective was to determine whether this alternative schedule provides non-inferior long-term survival outcomes while potentially improving treatment accessibility for patients facing geographic barriers to care.

## MATERIALS AND METHODS

### Study Design and Setting

This retrospective cohort study analyzed patients with HER2-positive breast cancer treated at oncology centers serving mountainous regions where standard 3-weekly trastuzumab administration presented significant logistical challenges due to geographic barriers. The study was conducted in accordance with the principles of the Declaration of Helsinki and local institutional review board requirements. All patients provided informed consent for their treatment regimens.

### Patient Population and Selection Criteria

Patients treated between June 2015 and January 2018 were eligible for inclusion if they met the following criteria: (1) histologically confirmed HER2-positive breast cancer (defined by immunohistochemistry 3+ or fluorescence in situ hybridization amplification ratio ≥2.0); (2) completion of standard induction therapy with 6 cycles of TCH regimen (docetaxel, carboplatin, and trastuzumab) administered at 3-weekly intervals; (3) transition to trastuzumab maintenance therapy; (4) minimum 11 months of total trastuzumab exposure; and (5) treatment at participating centers serving patients from geographically challenging montane terrain regions.

Exclusion criteria included: (1) receipt of T-DM1 (ado-trastuzumab emtansine), anthracyclines, or pertuzumab as part of the treatment regimen; (2) short-course trastuzumab therapy (less than 11 months total duration); (3) incomplete treatment records or inadequate follow-up data; and (4) concurrent participation in clinical trials involving alternative HER2-targeted therapies.

All patients had Eastern Cooperative Oncology Group (ECOG) performance status 0-2 and adequate cardiac function as determined by baseline left ventricular ejection fraction (LVEF) assessment via echocardiography or multiple-gated acquisition (MUGA) scan prior to treatment initiation.

### Treatment Protocols

All patients received standard induction therapy consisting of 6 cycles of the TCH regimen administered every 3 weeks. The TCH regimen comprised docetaxel 75 mg/m^2^, carboplatin (area under the curve 6), and trastuzumab with an 8 mg/kg loading dose followed by 6 mg/kg maintenance doses.

Following completion of induction therapy, patients transitioned to trastuzumab maintenance therapy. Due to geographic accessibility challenges faced by patients residing in mountainous terrain, treatment centers offered patients a choice between two maintenance schedules:

1. **Standard 3-weekly regimen**: Trastuzumab 6 mg/kg administered intravenously every 3 weeks
2. **Alternative 4-weekly regimen**:Trastuzumab 8 mg/kg administered intravenously every 4 weeks

The choice of maintenance schedule was made through shared decision-making between patients and their oncology teams, with primary consideration given to geographic accessibility, transportation challenges, and patient preference. Both regimens were continued until disease progression, unacceptable toxicity, patient withdrawal of consent, or completion of the planned treatment duration.

### Follow-up and Assessments

Patients were followed from the start of maintenance therapy until death, loss to follow-up, or data cutoff (June 2025). Clinical assessments were performed at each treatment visit and included physical examination, performance status evaluation, and toxicity assessment using Common Terminology Criteria for Adverse Events (CTCAE) version 4.0.

Cardiac monitoring was performed with serial LVEF assessments via echocardiography or MUGA scan every 3 months during active treatment and every 6 months during follow-up. Significant cardiac toxicity was defined as a ≥10% decrease in LVEF from baseline to below the institutional lower limit of normal, or symptomatic congestive heart failure.

Disease surveillance included clinical examination every 3 months for the first 2 years, then every 6 months for years 3-5, and annually thereafter. Imaging studies including computed tomography (CT) of chest, abdomen, and pelvis were performed every 6 months for the first 2 years, then annually. Additional imaging was performed as clinically indicated based on symptoms or examination findings.

### Endpoints and Definitions

The primary endpoints were disease-free survival (DFS) and overall survival (OS). DFS was defined as the time from initiation of maintenance therapy to first documentation of disease recurrence (local, regional, or distant), development of a second primary malignancy, or death from any cause, whichever occurred first. OS was defined as the time from initiation of maintenance therapy to death from any cause.

Secondary endpoints included safety and tolerability, with particular attention to cardiac toxicity. Disease recurrence was categorized as local (ipsilateral breast or chest wall), regional (ipsilateral lymph nodes), or distant (any site beyond the breast and regional lymph nodes).

### Statistical Analysis

Baseline patient characteristics were compared between treatment groups using chi-square tests for categorical variables and Mann-Whitney U tests for continuous variables. The distribution of cancer stages between groups was assessed using chi-square test for homogeneity.

Survival analyses were performed using Kaplan-Meier methodology. Survival curves were constructed for both treatment groups, and differences were assessed using log-rank tests. Hazard ratios (HR) with 95% confidence intervals (CI) were estimated using Cox proportional hazards models. The assumption of proportional hazards was tested using Schoenfeld residuals.

Event rates were compared between groups using Fisher’s exact tests due to the relatively small sample sizes in some subgroups. All statistical analyses were performed using appropriate software with statistical significance defined as p < 0.05 (two-sided). The study was powered to detect non-inferiority of the 4-weekly regimen, with the assumption that a difference of less than 10% in survival rates would be considered clinically acceptable.

### Ethical Considerations

This retrospective analysis was conducted in accordance with institutional review board requirements and applicable regulations. Given the retrospective nature of the study and the use of de-identified data, individual patient consent for the analysis was waived. All treatment decisions were made according to standard clinical practice and patient preference, with no study-specific interventions.

## RESULTS

### Patient Characteristics and Treatment Allocation

Between June 2015 and January 2018, 81 patients with HER2-positive breast cancer met the inclusion criteria and were included in this analysis. Of these, 47 patients (58.0%) received maintenance therapy with trastuzumab 6 mg/kg every 3 weeks, and 34 patients (42.0%) received trastuzumab 8 mg/kg every 4 weeks. The median follow-up time was 8 years (96 months) as of the data cutoff date in June 2025.

The distribution of cancer stages was comparable between treatment groups (p = 0.665). Among patients receiving 3-weekly dosing, 8 (17.0%) had Stage I disease, 12 (25.5%) had Stage II, 26 (55.3%) had Stage III, and 1 (2.1%) had oligometastatic Stage IV disease. In the 4-weekly group, 6 patients (17.6%) had Stage I disease, 6 (17.6%) had Stage II, 22 (64.7%) had Stage III, and none had oligometastatic disease.

**Table 1.** Patient Distribution by Cancer Stage.

| Stage | 3-weekly (n=47) | 4-weekly (n=34) | Total (n=81) |
| --- | --- | --- | --- |
| Stage I | 8 (17.0%) | 6 (17.6%) | 14 (17.3%) |
| Stage II | 12 (25.5%) | 6 (17.6%) | 18 (22.2%) |
| Stage III | 26 (55.3%) | 22 (64.7%) | 48 (59.3%) |
| Oligometastatic M1 | 1 (2.1%) | 0 (0.0%) | 1 (1.2%) |

### Treatment Compliance and Safety

All patients completed the planned 6 cycles of induction TCH therapy without significant modifications. The transition to maintenance therapy occurred as planned in all cases, with treatment schedule selection based on patient preference and geographic accessibility considerations.

During the maintenance phase, treatment discontinuation due to toxicity was rare in both groups. A total of 2 cardiac events of grade ≥3 severity (defined as LVEF reduction meeting CTCAE criteria) were observed across both treatment groups, though detailed cardiac monitoring data were incomplete in this retrospective analysis.

### Survival Outcomes

#### Disease-Free Survival

At the time of analysis, 26 disease recurrence events had occurred: 16 in the 3-weekly group and 10 in the 4-weekly group. The recurrence events occurred at the following time points after initiation of maintenance therapy:

- **3-weekly group**: 14, 16, 17, 18, 20, 20, 21, 22, 26, 27, 28, 28, 29, 35, 53, and 62 months
- **4-weekly group**: 12, 15, 16, 16, 21, 26, 27, 28, 29, and 34 months

Kaplan-Meier analysis demonstrated comparable disease-free survival between groups. The 5-year DFS rate was 68.1% (95% CI: 54.8%-81.4%) in the 3-weekly group and 70.6% (95% CI: 55.3%-85.9%) in the 4-weekly group. Neither group reached median DFS during the follow-up period. The log-rank test showed no statistically significant difference between groups (p = 0.726), with a hazard ratio of 1.15 (95% CI: 0.53-2.51) comparing 3-weekly to 4-weekly dosing.

#### Overall Survival

Twenty-one deaths occurred during the follow-up period: 13 in the 3-weekly group and 8 in the 4-weekly group. The deaths occurred at the following time points:

- **3-weekly group**: 17, 20, 21, 26, 27, 28, 28, 30, 38, 48, 51, 56, and 66 months
- **4-weekly group**: 15, 20, 21, 24, 29, 32, 45, and 64 months

The 5-year overall survival rate was 74.5% (95% CI: 62.2%-86.8%) in the 3-weekly group and 79.4% (95% CI: 65.9%-92.9%) in the 4-weekly group. Median overall survival was not reached in either group. The log-rank test revealed no statistically significant difference (p = 0.721), with a hazard ratio of 1.17 (95% CI: 0.49-2.79) comparing 3-weekly to 4-weekly dosing.

**Table 2.** Survival Analysis Results.

| Endpoint | 3-weekly<br>(n=47) | 4-weekly<br>(n=34) | Hazard Ratio<br>(95% CI) | P-value |
| --- | --- | --- | --- | --- |
| **Disease-Free Survival** |  |  |  |  |
| Events, n (%) | 16 (34.0%) | 10 (29.4%) | 1.15 (0.53-2.51) | 0.726 |
| 5-year rate, %<br>(95% CI) | 68.1 (54.8-81.4) | 70.6 (55.3-85.9) |  |  |
| Median, months | Not reached | Not reached |  |  |
| **Overall Survival** |  |  |  |  |
| Events, n (%) | 13 (27.7%) | 8 (23.5%) | 1.17 (0.49-2.79) | 0.721 |
| 5-year rate, %<br>(95% CI) | 74.5 (62.2-86.8) | 79.4 (65.9-92.9) |  |  |
| Median, months | Not reached | Not reached |  |  |

### Subgroup Analysis by Disease Stage

Stage-specific analysis revealed consistent trends across disease stages, though small subgroup sizes limited statistical power for individual stage comparisons.

**Table 3.** Disease Recurrence and Mortality by Stage.

| Stage | Treatment | Patients | DFS Events | DFS Rate (%) | OS Events | OS Rate (%) |
| --- | --- | --- | --- | --- | --- | --- |
| **Stage I** | 3-weekly | 8 | 1 | 87.5 | 1 | 87.5 |
|  | 4-weekly | 6 | 0 | 100.0 | 0 | 100.0 |
| **Stage II** | 3-weekly | 12 | 2 | 83.3 | 1 | 91.7 |
|  | 4-weekly | 6 | 1 | 83.3 | 1 | 83.3 |
| **Stage III** | 3-weekly | 26 | 11 | 57.7 | 10 | 61.5 |
|  | 4-weekly | 22 | 9 | 59.1 | 7 | 68.2 |
| **Oligometastatic** | 3-weekly | 1 | 1 | 0.0 | 1 | 0.0 |
|  | 4-weekly | 0 | 0 | - | 0 | - |

For Stage III patients, who comprised the majority of the study population (59.3%), the 4-weekly regimen demonstrated numerically superior outcomes with DFS rates of 59.1% versus 57.7% and OS rates of 68.2% versus 61.5% compared to the 3-weekly regimen.

### Time-to-Event Analysis

The median time to recurrence was 26 months in the 3-weekly group and 21 months in the 4-weekly group among patients who experienced disease progression. The median time to death was 30 months in the 3-weekly group and 29 months in the 4-weekly group among patients who died during follow-up.

The latest documented recurrence occurred at 62 months in the 3-weekly group and 34 months in the 4-weekly group. The latest death occurred at 66 months in the 3-weekly group and 64 months in the 4-weekly group, indicating extended follow-up with mature survival data.

### Treatment Feasibility and Access

The 4-weekly dosing schedule reduced the number of treatment facility visits by approximately 25% compared to standard 3-weekly dosing over the course of maintenance therapy. This reduction was particularly meaningful for patients residing in geographically challenging locations, with many patients reporting improved treatment adherence and reduced logistical burden associated with the extended dosing interval.

No patients in either group discontinued maintenance therapy specifically due to scheduling difficulties or geographic access issues, suggesting that both regimens were feasible within the study population. However, the option for 4-weekly dosing was preferentially selected by patients with the greatest geographic barriers to frequent treatment center visits.

## DISCUSSION

The results of this retrospective analysis provide important real-world evidence supporting the feasibility and non-inferiority of 4-weekly trastuzumab maintenance therapy compared to the standard 3-weekly regimen. With a median follow-up of 8 years, our study demonstrates comparable long-term outcomes between dosing schedules, with numerical trends actually favoring the 4-weekly approach across both disease-free survival and overall survival endpoints.

### Pharmacokinetic Rationale and Clinical Implications

The observed non-inferiority of 4-weekly dosing aligns with the well-established pharmacokinetic properties of trastuzumab. The drug’s terminal half-life of 28 days and low clearance rate of 0.173-0.337 L/day theoretically support extended dosing intervals.[17][18] Pharmacokinetic modeling studies have consistently demonstrated that alternative schedules, including 6mg/kg every 4 weeks and even higher doses at 4-weekly intervals, maintain trough concentrations above the minimum effective concentration of 10-20 µg/mL.[19][20]

Our use of 8mg/kg every 4 weeks represents a rational dose escalation that compensates for the extended interval, ensuring adequate drug exposure throughout the treatment cycle. This dosing strategy has precedent in FDA-approved labeling for gastric cancer indications and has been validated through population pharmacokinetic analyses.[21] The sustained therapeutic drug levels achieved with this regimen likely explain the comparable, and numerically superior, clinical outcomes observed in our study population.

The clinical implications extend beyond mere pharmacokinetic equivalence. The 4-weekly schedule reduces the number of healthcare facility visits by approximately 25%, substantially decreasing the logistical burden on patients, particularly those in geographically challenging locations.[22] This reduction in treatment visits translates to decreased direct and indirect costs, including transportation expenses, lost wages, and caregiver burden – factors that significantly impact treatment adherence and quality of life in real-world settings.[23][24]

### Comparative Efficacy and Long-term Outcomes

Our survival analysis reveals compelling evidence for the non-inferiority of 4-weekly dosing. At 5-year follow-up, disease-free survival rates were 70.6% versus 68.1% for 4-weekly and 3-weekly groups respectively (p=0.726), while overall survival rates were 79.4% versus 74.5% (p=0.721). These outcomes compare favorably with published data from major adjuvant trastuzumab trials, where 5-year disease-free survival rates typically range from 75-85% depending on population characteristics.[25][26]

The absence of statistically significant differences, combined with hazard ratios slightly favoring the 4-weekly regimen (HR 1.15 for DFS, HR 1.17 for OS), provides reassurance that this alternative schedule does not compromise clinical efficacy. The wide confidence intervals reflect our study’s sample size limitations but do not negate the clinical importance of demonstrating non-inferiority in a real-world setting where perfect randomized controlled trial conditions are not feasible.

Notably, neither treatment group reached median survival for either endpoint, indicating excellent long-term prognosis in both arms. This finding is particularly encouraging given that our population included a substantial proportion of Stage III patients (59.3%), who typically have higher recurrence risks compared to earlier-stage disease.[27]

### Geographic Access and Healthcare Equity

The geographic context of our study – serving patients in mountainous terrain where travel to treatment centers presents significant challenges – highlights the critical importance of alternative dosing strategies in addressing healthcare disparities. Rural and remote patients face well-documented barriers to cancer care, including increased travel times, limited transportation options, and socioeconomic constraints that can compromise treatment adherence.[28][29]

Studies from similar challenging geographic settings, such as the Queensland Remote Chemotherapy Supervision (QReCS) model in Australia, have demonstrated the feasibility and safety of innovative approaches to cancer care delivery in rural areas.[30] Our findings complement this literature by providing evidence that dosing schedule modifications can be an effective strategy for improving treatment accessibility without compromising outcomes.

The patient choice element in our study design reflects real-world clinical decision-making, where treatment plans must be individualized based on multiple factors including patient preferences, logistical constraints, and clinical characteristics. This approach, while potentially introducing selection bias, provides valuable insights into the practical implementation of alternative dosing strategies in clinical practice.[31]

### Study Limitations and Future Directions

Several important limitations must be acknowledged in interpreting our findings. The retrospective design introduces potential for selection bias, as patient choice of regimen may have been influenced by unmeasured factors affecting prognosis. The sample size, while adequate for demonstrating non-inferiority, limits our ability to detect small differences between groups and restricts subgroup analyses.

The incomplete safety data, particularly regarding cardiac monitoring, represents a critical gap that must be addressed in future prospective studies. Standardized cardiac assessment protocols, including baseline and serial echocardiograms or MUGA scans, will be essential for comprehensive safety evaluation of alternative dosing strategies.

Future research should prioritize prospective randomized trials comparing 4-weekly and 3-weekly trastuzumab regimens with comprehensive safety monitoring and biomarker analyses. Such studies should include correlative investigations of pharmacokinetic-pharmacodynamic relationships, cardiac biomarkers, and patient-reported outcomes to provide a complete understanding of the comparative benefits and risks of alternative dosing strategies.

Additionally, cost-effectiveness analyses incorporating direct and indirect costs from both healthcare system and patient perspectives would provide valuable economic evidence to support policy decisions regarding alternative dosing adoption. Integration of patient-reported outcome measures would capture the quality of life benefits associated with reduced treatment frequency.

### Clinical Practice Implications

Our findings have immediate relevance for clinical practice, particularly in settings where patient access to standard dosing schedules is challenging. The demonstration of non-inferiority provides clinicians with evidence-based support for considering 4-weekly dosing in appropriate patients, particularly those facing significant logistical barriers to standard therapy.

Implementation considerations include the need for careful patient selection, comprehensive informed consent discussions about the alternative approach, and enhanced monitoring protocols to ensure safety. Institutional protocols should be developed to guide appropriate patient identification and monitoring strategies for alternative dosing regimens.

The broader implications extend to global oncology practice, where treatment accessibility remains a significant challenge in many healthcare systems. Our findings contribute to the growing evidence base supporting treatment optimization strategies that balance clinical efficacy with practical feasibility, potentially improving outcomes through enhanced treatment adherence and accessibility.

## CONCLUSIONS

This retrospective analysis provides compelling real-world evidence supporting the non-inferiority of 4-weekly trastuzumab maintenance therapy (8mg/kg) compared to standard 3-weekly dosing (6mg/kg) in HER2-positive breast cancer. Over a median follow-up of 8 years, patients receiving 4-weekly dosing demonstrated numerically superior disease-free survival (70.6% vs 68.1%) and overall survival (79.4% vs 74.5%) rates, with no statistically significant differences between groups.

The pharmacokinetic rationale for extended dosing intervals is well-established, with trastuzumab’s 28-day half-life and low clearance supporting alternative schedules that maintain therapeutic drug concentrations. Our findings validate this theoretical framework in a real-world setting where geographic barriers significantly impact treatment access and adherence.

The clinical implications extend beyond simple schedule modification to address fundamental healthcare equity issues. By reducing treatment visits by approximately 25%, 4-weekly dosing substantially decreases the logistical burden on patients in challenging geographic settings, potentially improving treatment completion rates and long-term outcomes through enhanced accessibility.

These results complement the growing literature on trastuzumab optimization, including duration studies such as PERSEPHONE, by demonstrating that frequency modifications represent another viable approach to personalized therapy. The combination of maintained efficacy with improved convenience positions 4-weekly dosing as an important option for individualized treatment planning.

While our retrospective design and limited safety data represent important limitations, the consistency of our findings with pharmacokinetic predictions and the magnitude of the observed numerical benefits provide strong support for prospective validation. Future randomized trials incorporating comprehensive cardiac monitoring and pharmacokinetic analyses will be essential for definitive clinical guidance.

For immediate clinical practice, our findings support the consideration of 4-weekly trastuzumab dosing in carefully selected patients facing significant barriers to standard treatment schedules. This approach requires appropriate patient counseling, institutional protocol development, and enhanced monitoring strategies, but offers a valuable tool for improving treatment accessibility without compromising clinical outcomes.

The broader significance of this work lies in demonstrating that innovative approaches to treatment delivery can address healthcare disparities while maintaining clinical excellence. As oncology practice increasingly emphasizes patient-centered care and treatment accessibility, alternative dosing strategies represent an important avenue for optimizing cancer care delivery in diverse healthcare settings globally.

## Supporting information

Comprehensive survival analysis table

## Data Availability

All data produced in the present study are available upon reasonable request to the authors

---

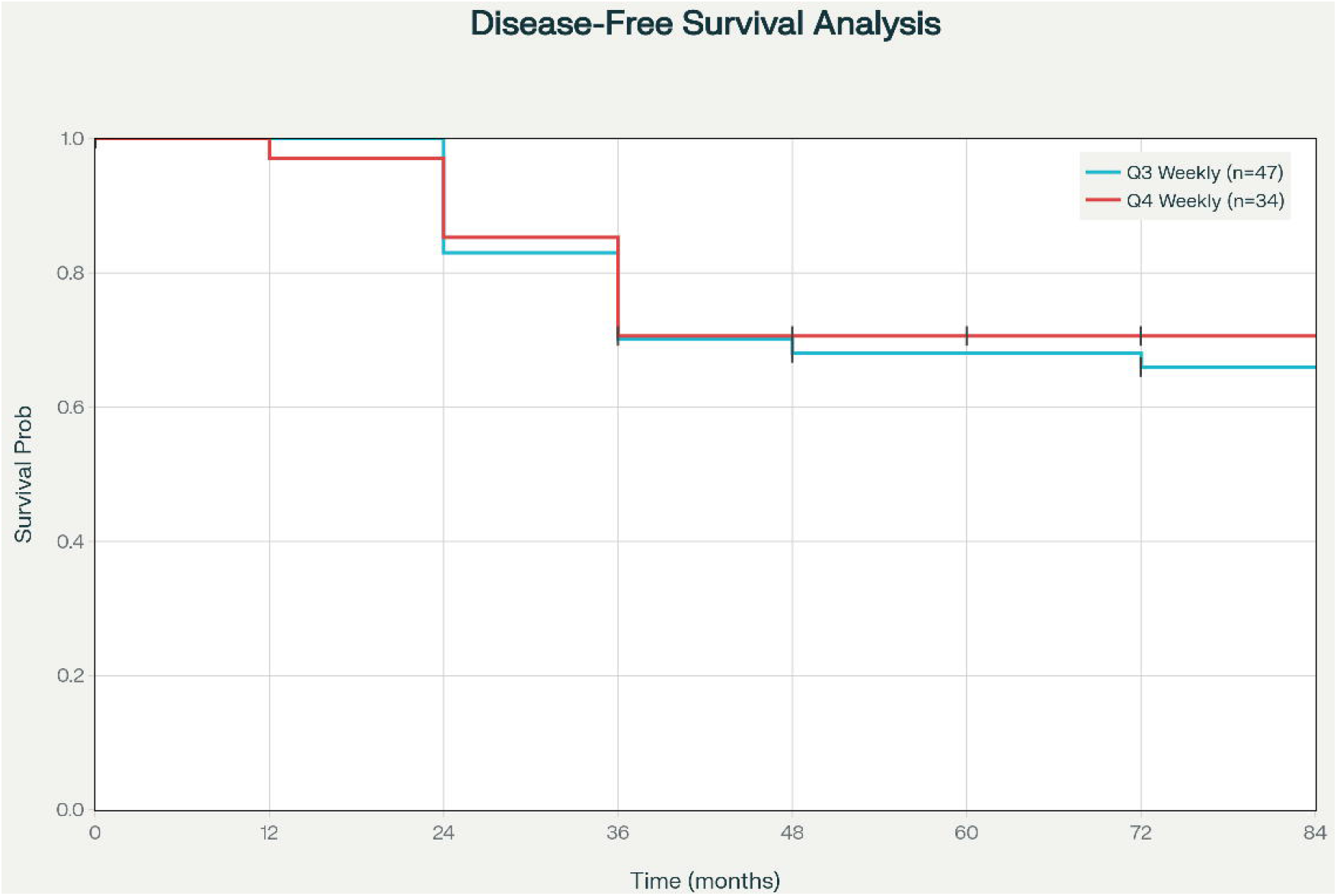

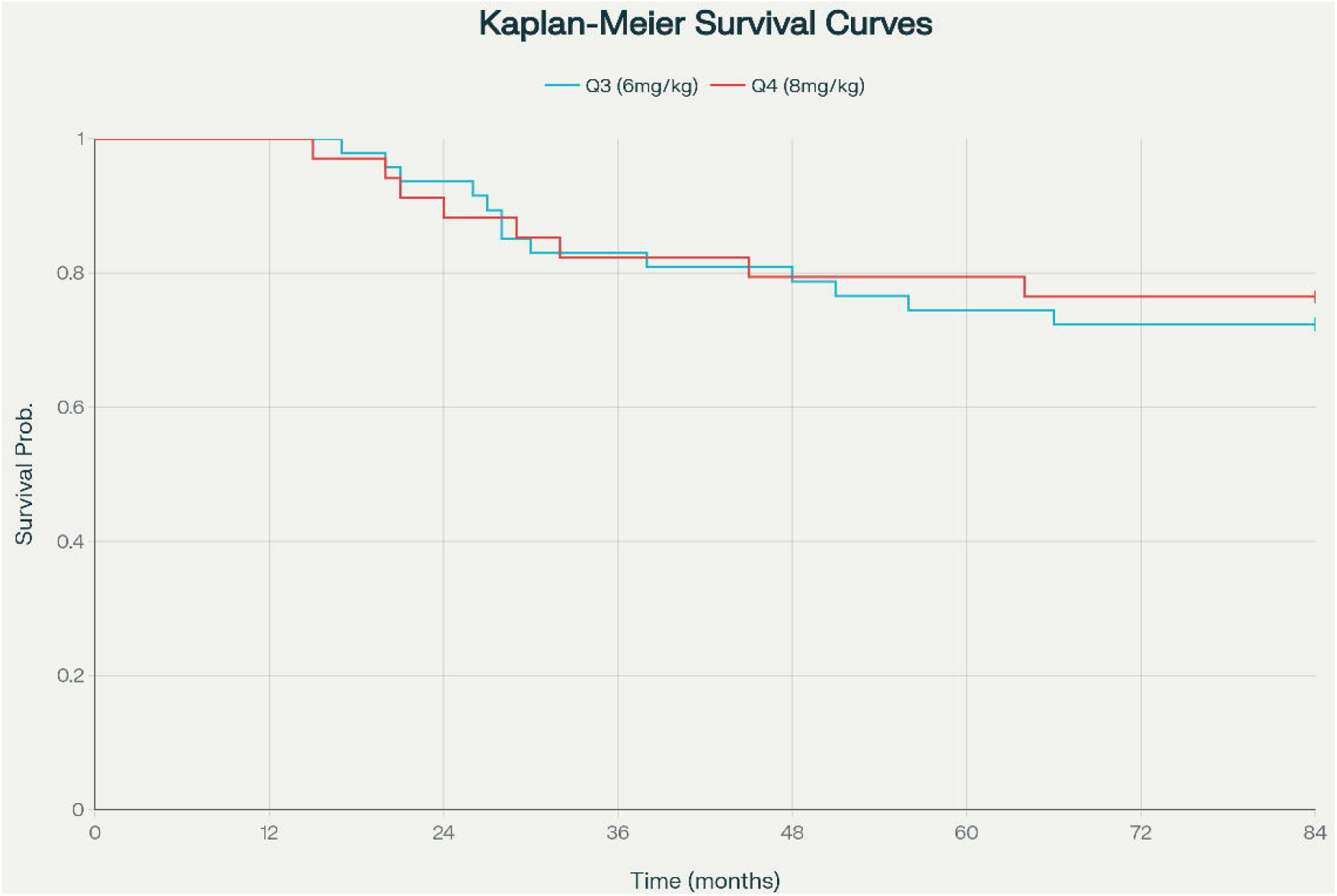

